# The confounded crude case-fatality rates (CFR) for COVID-19 hide more than they reveal - a comparison of age-specific and age-adjusted CFRs between six countries

**DOI:** 10.1101/2020.05.09.20096503

**Authors:** Manfred S Green, Victoria Peer, Naama Swartz, Dorit Nitzan

**Affiliations:** School of Public Health, University of Haifa, Israel; World Health Organization, European Region, Copenhagen, Denmark

**Keywords:** COVID-19, case-fatality rates, age-specific rates, age-adjusted rates, confounding

## Abstract

**Background:** The reported crude case-fatality rates (CFRs) vary widely between countries. The serious limitations of using crude COVID-19 CFRs for comparisons between countries have been addressed in the literature but are often overlooked or misunderstood, both in the scientific literature and in the media. In this paper we examined the extent to which age distribution of the cases is responsible for the differences in CFRs between countries.

**Methods:** Data on COVID-19 were extracted from the reports of individual countries. Overall and age-specific CFRs were available for six countries. The CFRs by country were adjusted for age using the direct method, using the combined age-specific number of cases of all six countries as the standard population.

**Findings:** The age distribution of the cases varied widely between countries. The crude CFRs varied between 1.6% and 11%. The differences in the age-specific CFRs were much smaller and the age-adjusted rates were much closer than the crude rates. The ratio of the crude CFR for the country with the highest CFR to that with the lowest, was reduced substantially for the age-adjusted rates, from 7.4 to 2.3

**Conclusions:** The age structure of the cases dramatically impacts on the differences in the crude CFRs between countries. Adjusting for age substantially reduces this variation. Other factors such as the differences in the definition of the denominators, the definition of a case and the standard of healthcare are likely to account for much of the residual variation. It is misleading to compare the crude COVID-19 CFRs between countries and should be avoided. At the very least, comparisons should be based on age-specific and age-adjusted rates.

## Background

The novel coronavirus SARS-2-CoV 2019 was first identified in Wuhan in China in early December 2019. On January 30, 2020, the World Health Organization declared the disease caused by the virus, a Public Health Emergency of International Concern (PHEIC) (1) and on March 11, 2020, it was declared as a pandemic. By May, 2020, almost all countries had been affected with more than three million cases of COVID-19 reported. Early estimates indicated that the average proportion of deaths among the diagnosed cases, defined as the case-fatality rate (CFR), was around 2.3% (2). However, subsequently, the reported crude COVID-19 CFRs varied widely between countries (3,4). A related concept is the infection fatality rate (IFR) which includes asymptomatic cases in the denominator, which need to be identified by screening tests. Since the IFR is rarely available for COVID-19, in this paper we consider only the CFR. The limitations of comparing crude CFRs in general, has been evaluated previously (5). The strong positive association between the COVID-19 CFR and age has been demonstrated both as in observational studies (6) and in a model-based analysis (7), particularly over the age of 40.

The interpretation of the CFR depends on the context in which it is used. In a single cohort of patients, it can indicate the severity of the disease at a single point in time. It can also be used to assess trends in the impact of changes in health care over time. In the current context of the COVID-19 pandemic, CFRs are commonly compared between countries, which may lead to speculation on differences in healthcare. In other words, it may be concluded that the CFRs somehow reflect the successes and failures of the different countries to deal with serious cases. For diseases in general and for COVID-19 in particular, a number of factors could impact on the both the numerator and denominator of the CFR. Examples are possible misclassification of causes of death and the variations in the definition of cases in the denominator. In this paper, we examined the contribution of the age distribution of the cases when comparing the COVID-19 CFRs between different countries. We focused our analyses on six countries, with widely varying COVID-19 CFRs.

## Methods

We studied published crude and age-specific CFRs in cohorts of cases of COVID-19 in six countries, with varying periods of follow-up. The countries chosen were based on the accessibility of the data at the beginning of May, 2020. The data for each country were not necessarily updated to the time of the study, and the CFR’s may have changed over time. Age-specific data on the cases and deaths were available for six countries: Italy (8), Spain (9), Sweden (10), China (11), S Korea (12) and Israel (unpublished data). All published data on cases and deaths by ten-year age groups (0-9, 10-19, …..80+). The outcome variable was defined as the crude CFR defined as the number of deaths divided by the number of reported cases. The exposure variable was the individual country. Age-group was considered as a confounding variable. Age-adjustment was carried by the direct method, using the distribution of the combined cases of all six countries by age group as the standard population. 95% confidence intervals were computed for each age-adjusted rate using WinPepi (PEPI-for-Windows).

## Ethical considerations

Open access aggregative and anonymous data were used and there was no need for ethics committee approval.

## Results

The age-specific number of cases, number of deaths and the crude CFRs by country are given in Table 1. They distributions vary markedly. The distribution of the cases for Israel and South Korea are heavily weighted in the 20-39 age group, China has a more balanced distribution and Italy and Sweden are heavily weighted in the over 70 age groups.

**Table 1.** Cases, deaths and CFRs by age group.

| **Age group** | **Country** | **Cases** | **Died** | **%CFR** |
| --- | --- | --- | --- | --- |
| 0-39 | China | 12,184 | 26 | 0.21% |
|  | Israel | 9,457 | 3 | 0.03% |
|  | Italy | 32,196 | 66 | 0.20% |
|  | S-Korea | 4,814 | 2 | 0.04% |
|  | Spain | 38,614 | 93 | 0.24% |
|  | Sweden | 6,271 | 24 | 0.38% |
| 40-49 | China | 8,571 | 38 | 0.44% |
|  | Israel | 1,966 | 3 | 0.15% |
|  | Italy | 27,553 | 246 | 0.89% |
|  | S-Korea | 1,421 | 3 | 0.21% |
|  | Spain | 35,135 | 201 | 0.57% |
|  | Sweden | 4,107 | 36 | 0.88% |
| 50-59 | China | 10,008 | 130 | 1.30% |
|  | Israel | 1,904 | 9 | 0.47% |
|  | Italy | 38,399 | 993 | 2.59% |
|  | S-Korea | 1,953 | 15 | 0.77% |
|  | Spain | 42,794 | 611 | 1.43% |
|  | Sweden | 5,198 | 112 | 2.15% |
| 60-69 | China | 8,583 | 309 | 3.60% |
|  | Israel | 1,454 | 26 | 1.79% |
|  | Italy | 29,252 | 2,976 | 10.17% |
|  | S-Korea | 1,347 | 35 | 2.60% |
|  | Spain | 34,360 | 1,695 | 4.93% |
|  | Sweden | 3,608 | 263 | 7.29% |
| 70-79 | China | 3,918 | 312 | 7.96% |
|  | Israel | 843 | 66 | 7.83% |
|  | Italy | 31,627 | 7,849 | 24.82% |
|  | S-Korea | 708 | 72 | 10.17% |
|  | Spain | 32,443 | 4,632 | 14.28% |
|  | Sweden | 3,422 | 815 | 23.82% |
| 80+ | China | 1,408 | 208 | 14.77% |
|  | Israel | 668 | 177 | 26.50% |
|  | Italy | 55,020 | 15,825 | 28.76% |
|  | S-Korea | 485 | 115 | 23.71% |
|  | Spain | 55,779 | 11,954 | 21.43% |
|  | Sweden | 6,596 | 2,396 | 36.33% |

The distributions of the cases for each country are shown in Figure 1. It is clear that distributions vary widely and are not necessarily related to the age distribution of the population of the country. For example, for South Korea, there is a relatively large number of cases in the age group 20-29, due to an outbreak affecting that age group in particular.

**Figure 1.**
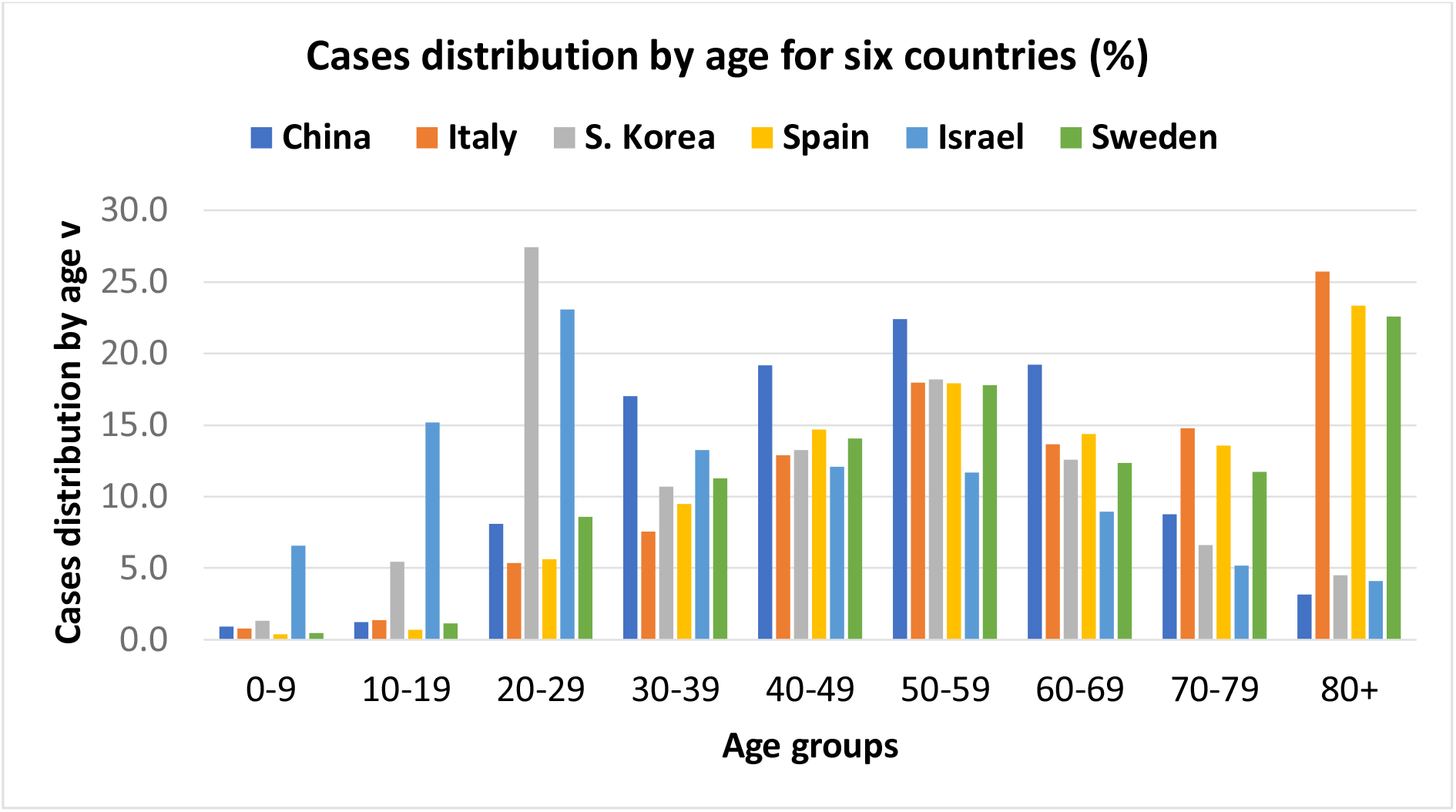
Cases distribution by age for six countries (China, Italy, S Korea, Spain, Israel and Sweden)

The age-specific CFRs are shown in Figure 2. While there are differences in the age-specific CFRs, the trend of steeply increasing CFRs in the oldest age groups is evident.

**Figure 2.**
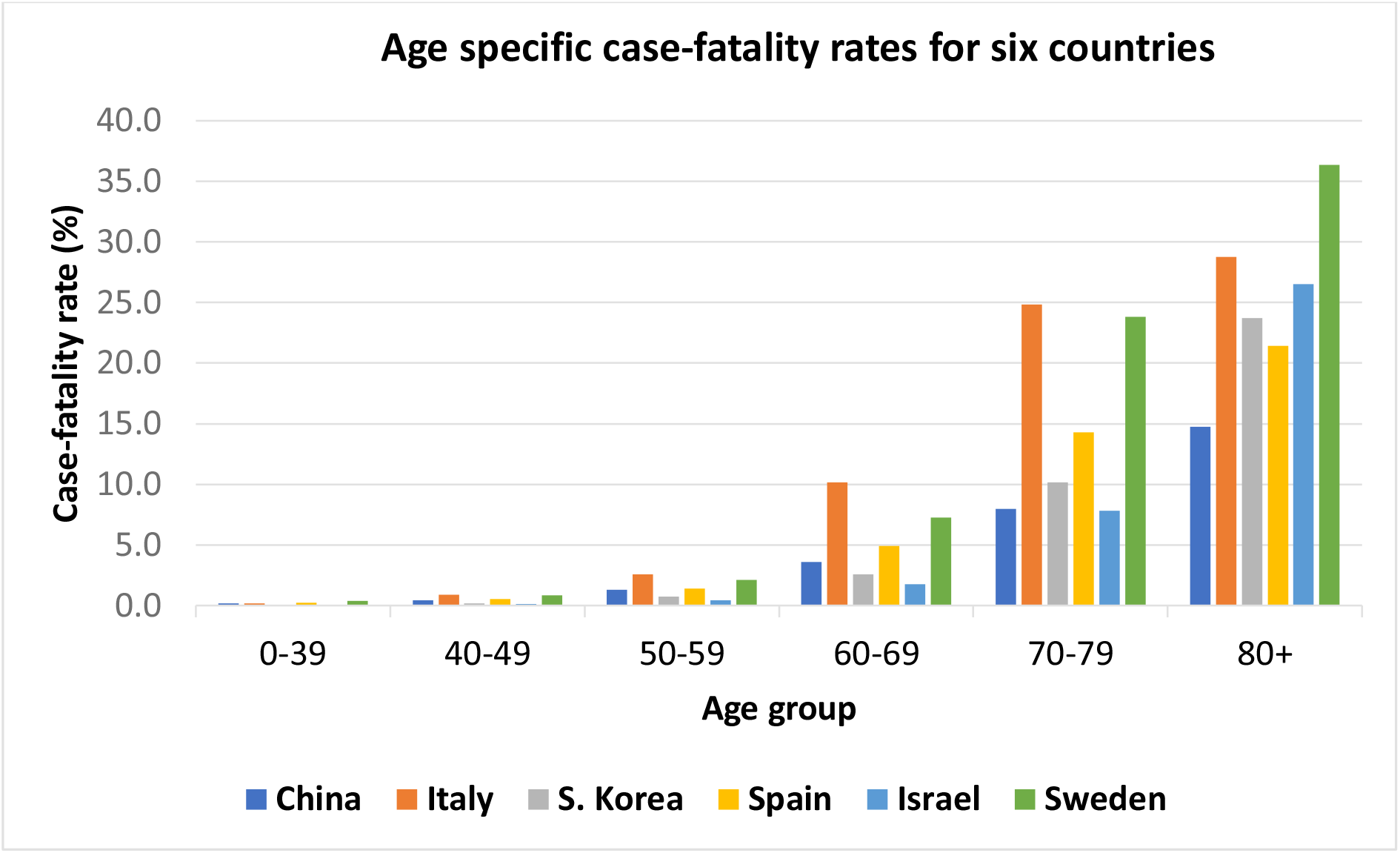
Age specific case-fatality rates for six countries – China, Italy, S Korea, Spain, Israel and Sweden.

The crude and age-adjusted CFRs are compared in Table 2. The crude CFRs varied from 1.7% for Israel to 13.1% for Italy and the ratios of the crude CFRs compared with lowest CFR, varied between 1.4 and 7.7. The age-adjusted CFRs varied between 5.1 % for China to 12.6% for Sweden and the ratios of the age-adjusted CFRs for each country compared with the lowest CFR adjusted varied between 1.4 to 2.5.

**Table 2.** Crude and age standardized case-fatality rates (CFR) for COVID-19 in six countries.

| Country | Crude CFR % | Age-adjusted CFR rate % (95% CI) | Ratio to lowest crude CFR | Ratio to lowest age-adjusted CFR |
| --- | --- | --- | --- | --- |
| Israel | 1.7 | 7.1 (6.4-7.9) | 1.0 | 1.4 |
| Korea | 2.3 | 7.0 (6.1-7.9) | 1.4 | 1.4 |
| China | 2.3 | 5.1 (4.7-5.5) | 1.4 | 1.0 |
| Spain | 8 | 7.6 (7.5-7.7) | 4.7 | 1.5 |
| Sweden | 12.5 | 12.6 (12.3-13.0) | 7.4 | 2.5 |
| Italy | 13.1 | 11.6 (11.5-11.7) | 7.7 | 2.3 |

Figure 3 shows graphically the marked reduction in the differences between the crude and age-adjusted CFRs for the six countries.

**Figure 3.**
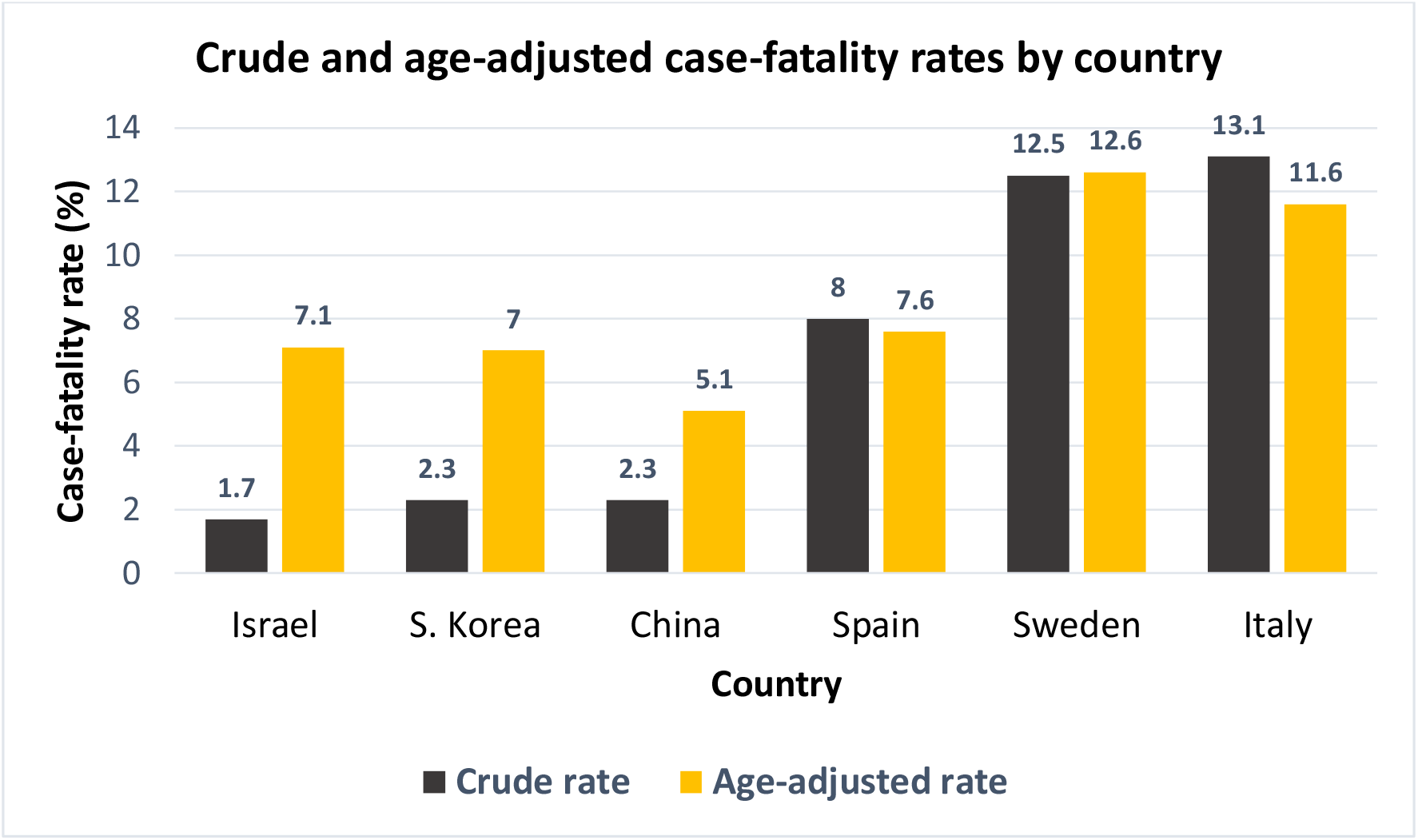
Crude and age-adjusted case-fatality rates by country for six countries.

## Meta-analyses by age group

We used meta-analytic methods to obtain weighted pooled estimates of the CFR’s by age group. The results of the meta-analysis are presented in the forest plot in Figure 4, which shows the CFRs by age group and country and the pooled CFR’s for each age group. There was considerable variation in the CFR’s by country within each age group, but the trends by age were very similar, increasing steeply after age 60. The pooled CFR’s were 0.17% in the age group 0-39, 0.52% for 40-49, 1.46% for 50-59, 5.07 for 60-69, 14.84% for 70-79, and 25.3% for 80+.

**Figure 4:**
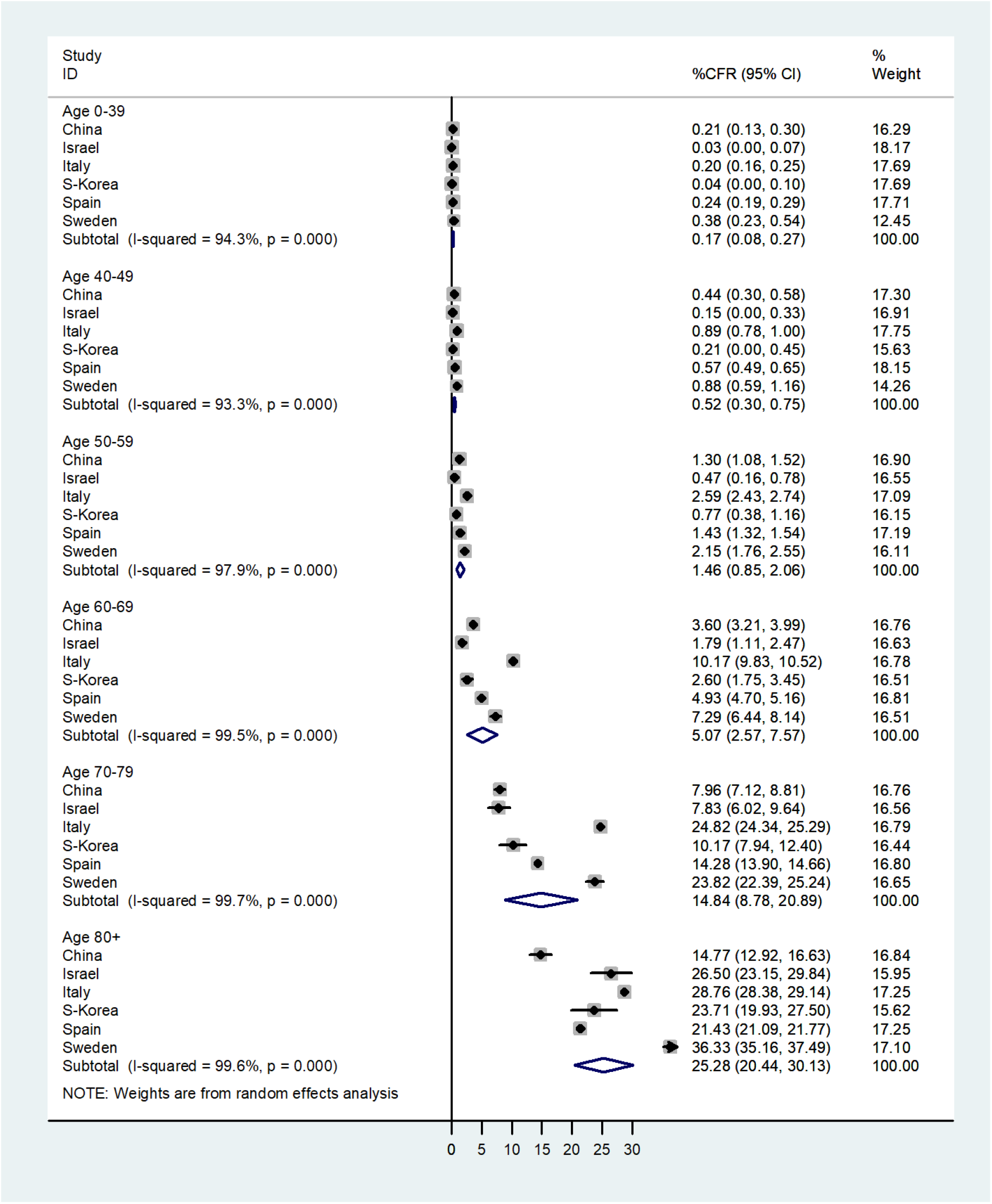
Forest plot of case-fatality rates by age group and country.

## Discussion

In our study, we examined crude, age-specific and age-adjusted CFRs for COVID-19 in six countries, with widely varying crude CFRs. The trends in the age-specific CFRs were remarkably similar in the six countries, with the CFR’s increasing steeply in those over 70. After adjusting for age, the marked differences in the crude CFRs were substantially reduced. These findings demonstrate the importance of accounting for age when comparing rates in general and CFRs in particular. The results of this study are strengthened by the use of national data or large datasets from a number of countries, with considerable differences in the extent of the pandemic in each country. It should be stressed that the age distribution of the cases was used to compute the age-adjusted CFRs and not the age distribution of the total population in each country.

In addition to the age distribution of the cases, the use of the CFR for comparisons between countries has other important limitations. Selection bias is clearly present when calculating the denominator on the basis of reported cases. As mentioned, the CFR must be distinguished from the IFR, which includes asymptomatic cases identified by deliberate or incidental screening with diagnostic tests. In addition, if only those with more severe symptoms are tested this will affect the denominator of the CFR and will depend on the testing strategy of each country. If more mild cases are identified, this is likely to reduce the CFR. Since there is a lag time between the reporting of the case and the death which can occur up to weeks later. In the country reports, cases and deaths are usually reported at the same time, so the cases in the denominator are usually an overestimate of the true denominator which should be the number of cases reported sometime earlier (13,14). This will have a more dramatic effect when the number of cases are rising rapidly. Selection bias may also affect the numerator if only deaths occurring in hospital are reported. Information bias can be present in both the numerator and denominator of the CFR. The definition of the cases due to may be biased due to the variability of the sensitivity and specificity of the diagnosis of COVID-19. Information bias in the numerator can occur when the cause of death is coded. This could be particularly problematic in elderly people with multiple co-morbidities.

However, while these factors will affect the calculation of the true CFR, the purpose of this paper is to demonstrate the dramatic effect of confounding by the age distribution of the cases when using crude CFRs for country comparisons. This was shown in an earlier paper when comparing three countries, and we have extended it to a comparison of six countries with widely different CFRs. In this study, the age structures of the population of the six countries included varied markedly. The percentage of the population 65 and over is 11% in Israel, in Italy 23% in Italy, S Korea 14%, in Spain 19%, in Sweden 20% and in China 11% (15). However, the main impact of confounding by age was due to the differences in the age distribution of the cases. This was largely due to the specific circumstances of exposure. For example, most of the cases in Italy occurred in an area of a particularly old population (3). In some countries, many of the cases were medical personnel, a large number of whom were relatively young women (16). In South Korea, a large percentage of the cases were young women associated with a specific religious group (17). In Germany, many of the cases were relatively young people returning from skiing holidays in Austria and Italy (18). In Israel, the largest outbreaks occurred in the ultra-orthodox Jewish community, where the number of children per family is much higher than in the general population. Other factors affecting the age distribution of the cases, depended on the frequency of outbreaks in homes for the elderly (19).

The results of this study once again demonstrate the pitfalls of comparing unadjusted rates. The assumption that differences between countries in testing policies or standard of treatment accounted for the wide discrepancies in CFRs, is not well-founded. This does not mean that there are no differences. For example, it is possible that where the health services were overloaded, younger patients were more likely to be admitted to intensive care units with better chances of survival. Clearly, the data are incomplete and other factors affecting CFRs such as case definitions, use of different denominators, underlying health conditions and the standard of health services are likely to play important roles. In order to assess the impact of these factors, age-specific and age-adjusted CFRs must be used.

## Data Availability

All the data in the study are publically available

## Conclusions

In addition to the selection and information biases inherent in computing CFRs, the age structure of the cases dramatically impacts on the differences in the crude CFRs between countries. Failure to account for this source of confounding markedly distorts the country comparisons. The substantial reduction in the differences in the age-adjusted CFRs suggest that differences in the standard of healthcare between these countries may not play as important a role in affecting the death rates, as some have hypothesized. Crude COVID-19 CFRs have no real use for between country comparisons and should be avoided. In general, for comparisons between groups and countries, age-adjusted CFRs can be used, but age-specific COVID-19 CFRs are generally far more meaningful.

## Acknowledgements

We express our appreciation to the official institutions of all countries for the providing their data on Covid-19.

